# COVID-19 containment in the Caribbean: the experience of Small Island Developing States

**DOI:** 10.1101/2020.05.27.20114538

**Authors:** MM Murphy, SM Jeyaseelan, C Howitt, N Greaves, H Harewood, KR Quimby, N Sobers, RC Landis, KD Rocke, IR Hambleton

**Author notes:** **Address for Correspondence**. Ian Hambleton.

## Abstract

**Background:** Small island developing states (SIDS) have limited absolute resources for responding to national disasters, including health emergencies. Since the first confirmed case of COVID-19 in the Caribbean on 1st March 2020, non-pharmaceutical interventions (NPIs) have been widely used to control the resulting COVID-19 outbreak. We document the variety of government measures introduced across the Caribbean and explore their impact on aspects of outbreak control.

**Methods:** Drawing on publically available information, we present confirmed cases and confirmed deaths to describe the extent of the Caribbean outbreak. We document the range of outbreak containment measures implemented by national Governments, focussing on measures to control movement and gatherings. We explore the temporal association of containment measures with the start of the outbreak in each country, and with aggregated information on human movement, using smartphone positioning data. We include a set of comparator countries to provide an international context.

**Results:** As of 25th May, the Caribbean reported 18,755 confirmed cases and 631 deaths. There have been broad similarities but also variation in the number, the type, the intensity, and particularly the timing of the NPIs introduced across the Caribbean. On average, Caribbean governments began controlling movement into countries 27 days before their first confirmed case and 23 days before comparator countries. Controls on movement within country were introduced 9 days after the first case and 36 days before comparators. Controls on gatherings were implemented 1 day before the first confirmed case and 30 days before comparators. Confirmed case growth rates and numbers of deaths have remained low across much the Caribbean. Stringent Caribbean curfews and stay-at-home orders coincided with large reductions in community mobility, regularly above 60%, and higher than most international comparator countries.

**Conclusion:** Stringent controls to limit movement, and specifically the early timing of those controls has had an important impact on containing the spread of COVID-19 across much of the Caribbean. Very early controls to limit movement into countries may well be particularly effective for small island developing states. With much of the region economically reliant on international tourism, and with steps to open borders now being considered, it is critical that the region draws on a solid evidence-base to balance the competing demands of economics and public health.

## INTRODUCTION

One in five members of the United Nations (UN) are small island developing states (SIDS); 38 countries with a combined population of around 61 million. (1) The majority of SIDS are in the Caribbean and Pacific, and in addition to common social, economic and environmental vulnerabilities they share limitations related to healthcare provision for rapidly aging populations with high burdens of non-communicable disease. (2-4) In the Caribbean, there are 16 UN recognised SIDS, with a further 13 island territories without UN status and with formal ties to extra-regional UN members (USA, UK, France, Netherlands). Despite this variation in geo-political affiliations, one regional body, the Caribbean Community (CARICOM), includes 20 of the 29 Caribbean countries and territories as members. Serving a combined population of around 16 million people, CARICOM represents the dominant structure for regional cooperation on economic, political, health and disaster response. (5, 6) Although many of the island states in the Caribbean are classified as high or middle income – a classification that reduces the available international support - there is now global recognition that SIDS represent a further vulnerable country grouping due to specific economic and climate change disadvantages. (7) They have limited absolute resources to systematically tackle the complexities of their national health burdens, including responding to acute health emergencies.

On March 1^st^ 2020 the first confirmed case of COVID-19 in the Caribbean, an Italian tourist, was reported in the Dominican Republic; one month after the first case in Italy, and three months after patient zero in China. (8, 9) By that time, 87 thousand cases in 59 countries had been confirmed, and the Caribbean region, whose economies are heavily dependent on tourist arrivals from Europe and North America, was on high alert. On March 11^th^ 2020, the World Health Organization declared a global pandemic. As of May 25 2020, there were 5.3 million confirmed cases worldwide, including 2.4 million cases in the Americas. (10) Since March, CARICOM had been actively developing regional public health responses to the COVID-19 pandemic. (11) At the time of the first identified cases among CARICOM member states, the regional response was in its infancy, and CARICOM members were also relying on local expertise and international evidence.

Implemented measures can be broadly classed as non-pharmaceutical interventions (NPIs). Globally, NPIs are typically introduced as public health responses to outbreaks, and in the case of COVID-19 have been the main method of outbreak control due to the lack of vaccine or pharmaceutical treatment options. (12) These containment measures are expected to slow the spread of the virus and reduce the severity of the epidemic peak by reducing physical contact, which in turn can reduce disease transmission, and has an ultimate goal of keeping healthcare demand below health system capacity.

While there have been many similarities in the decisions by CARICOM countries to quickly implement NPIs, there has been distinct variation in the number, the type, the intensity, and particularly the timing of the NPIs. Here we compare national responses across the Caribbean (20 CARICOM and two nonmember countries) and explore the potential impact of implemented NPIs. We focus on NPIs affecting human movement. In particular, we examine policies related to movement into countries, movement within countries, and control of mass gatherings, against the dynamics of confirmed cases, confirmed deaths, outbreak growth rates, and population mobility. Understanding how combinations and timing of NPIs work, and in which contexts, can inform the continued response to COVID-19 as well as future virulent outbreaks within SIDS.

## METHODS

Our main goal was to present a COVID-19 situation analysis for the Caribbean region during the initial outbreak period (April and May 2020). This period broadly represents the time before governments in the Caribbean began to gently ease their national containment measures. We present confirmed cases and confirmed deaths to describe the extent of the outbreak across the Caribbean. We document the range of containment measures implemented by Governments and explore the time between the start of the outbreak in each country and the start of containment. We describe the temporal association of key containment measures and aggregated information on human movement, using smartphone positioning data.

### Data Sources

We drew on four data sources for this report. To describe the NPIs implemented in our included countries we used the ACAPS COVID-19 government measures database. This database collates measures implemented by governments worldwide in response to the coronavirus pandemic. It uses a variety of international, national, and media sources and includes a secondary review of collated information. Measures are grouped into 5 broad categories: movement restriction, social distancing, public health measures, governance and socioeconomic measures, and lockdown. (13) Information for six of the CARICOM counties, the United Kingdom overseas territories (or UKOTS: Anguilla, Bermuda, British Virgin Islands, Cayman Islands, Montserrat, Turks and Caicos Islands), were not included in the ACAPS database. To complete the CARICOM NPI information, and to generally ensure database accuracy, we systematically searched for missing or inaccurate NPI entries (see supplement for search protocol). To report the numbers of confirmed cases and deaths, we drew on two data sources. Primarily, we used the European Centre for Disease Control (ECDC) database for the distribution of confirmed COVID-19 cases and deaths worldwide. (14) We supplemented the ECDC numbers using data available each evening from the Johns Hopkins Coronavirus Resource Center. (15) To describe movement of people within their communities we used the Google-generated COVID-19 Community Mobility Reports. (16, 17) For selected countries (8 Caribbean countries, 8 comparator countries), these data describe the change in the number and length of visits to various grouped locations (grocery and pharmacy, parks, transit stations, retail and recreation, residential, and workplaces). The percentage change in movement for each day is compared to average movement on the same day of the week between 3rd Jan and 6th Feb 2020. Due to privacy concerns (if somewhere isn’t busy enough to ensure anonymity), data from the smaller countries of the Caribbean are not publicly available. Data are only collected from users who have opted-in to Google data collection, so the movement data are indicative not representative of a country’s population movement.

#### Included countries and territories

Our Caribbean surveillance work during the COVID-19 outbreak has centred on the 15 CARICOM member states and 5 associate members (See Table 1). We included all 20 members in this review and included 2 further Caribbean islands that have experienced major COVID-19 outbreaks: Cuba and Dominican Republic. Although Belize (Central America) and Guyana and Suriname (north-eastern South America) are geographically separated from the Caribbean islands they each have strong historical and socio-political links to the Caribbean island states. As full members of CARICOM each of these three territories are integrated into the Caribbean political, social, and economic reality. (18) We included 9 comparator countries from outside of the Caribbean region, which together represent a range of COVID-19 outbreak responses, particularly in terms of response timing and response stringency. Our comparator countries are Germany, Iceland, Italy, New Zealand, Singapore South Korea, Sweden, United Kingdom, Vietnam. In choosing our exemplar countries, we were careful to include examples of good national COVID-19 control (New Zealand, Singapore, South Korea, Vietnam), countries with a range of outbreak responses in Europe (Germany, Iceland, Italy, Sweden, United Kingdom), and island nations with small populations (New Zealand, Iceland, Singapore).

#### Statistical Methods

Our analyses are descriptive. We explored the extent of the Caribbean outbreak in two ways. First, we plotted the cumulative cases and deaths across the Caribbean as of 25-May-2020, stratifying into CARICOM and non-CARICOM states. Second, we calculated the growth rate for confirmed cases in each country by using the logarithm of the new daily cases then plotted a 7-day smoothed average growth rate over time for each country on a heatmap. We described the NPIs implemented in each country, grouping measures into those controlling movement into the country (border controls, and border closures), those controlling movement within a country (mobility restrictions, curfews, lockdown), and those controlling gatherings (limiting public gatherings, closing public services, and closing schools). These NPI groups are described in more detail in Box 1. Last, we explored the temporal association of containment measures with outbreak data and with movement data. For each country and for each broad containment group (controlling movement into a country, controlling movement within a country, controlling gatherings), we plotted the number of days between the date of first case and the date of the containment measure. Using Google data on community movements, we plotted the daily movement reduction (see Supplement) and the maximum average weekly movement reduction achieved by each country with available data, linking this timing with the implementation of two key containment measures associated with human movement control (curfews, lockdowns).

###### Box 1. Taxonomy of government-initiated non-pharmaceutical interventions (NPIs) ^1^

| Category | Measure | Description of Measure |
| --- | --- | --- |
| **Control Movement into Country** |  |  |
|  | Border controls | Any measure to intensify border controls, including health checks at border, visa restrictions or suspensions, and requirements for additional health documents. |
|  | Border closure: full | All points of entry closed including complete suspension of international flights and/or full airport closure |
|  | Border closure: partial | Some points of entry into country closed and/or passengers from certain destinations not permitted entry. This also includes any flight suspensions from specific destinations |
| **Control Movement in Country** |  |  |
|  | Mobility restrictions | Includes domestic travel restrictions with or without structured or ad-hoc security checks. |
|  | Curfews | Government order for people to remain in their home between specified hours (mostly at night). |
|  | Lockdown / stay-at-home order: full | 24-hour curfew and/or country under emergency “*stay at home*” order, and closure of public spaces. Only movement of essential workers allowed. Only essential services open. |
|  | Lockdown / stay-at-home order: partial | As full lockdown, except that some public spaces remain open and/or specific businesses remain open, in addition to essential services. |
| **Control Gatherings** |  |  |
|  | Limit public gatherings | Any measure to ban or reduce the number of people allowed at public gatherings (such as weddings, funerals, religious worship) and social occasions. |
|  | Close businesses or public services | Any measure to close or limit public access to non-healthcare public services, and/or private businesses. |
|  | Close schools | Any measure to close an educational facility, including tertiary education |

## RESULTS

In Table 1 we present the numbers of confirmed cases and deaths in each included country, along with selected country characteristics. The 22 Caribbean SIDS are generally geographically small, with small populations. In 2020, only 5 Caribbean SIDS have estimated populations over 1 million people (Trinidad and Tobago, Jamaica, Dominican Republic, Cuba, Haiti), and 9 have populations less than 100,000. Fourteen SIDS have land areas less than 100 square kms, leading to high population densities; 15 SIDS have densities in excess of 200 people per square km, against a global average of 59 per square km, and several Caribbean SIDS are amongst the most densely populated countries in the world (Haiti, Barbados, Bermuda). Although the United Nations Human Development Index (HDI) across the Caribbean is generally categorized as “High” (between 0.7 and 0.8), the average expenditure on health is around 6% (5.5% excluding Cuba), compared to a European average of 10%. Crucially, all Caribbean SIDS have a Global Health Security Index (GHS) below 40 (average 32, range 24 to 38) against a global average of 40.2 and an average among high-income nations of 51.9.

**Table 1.**
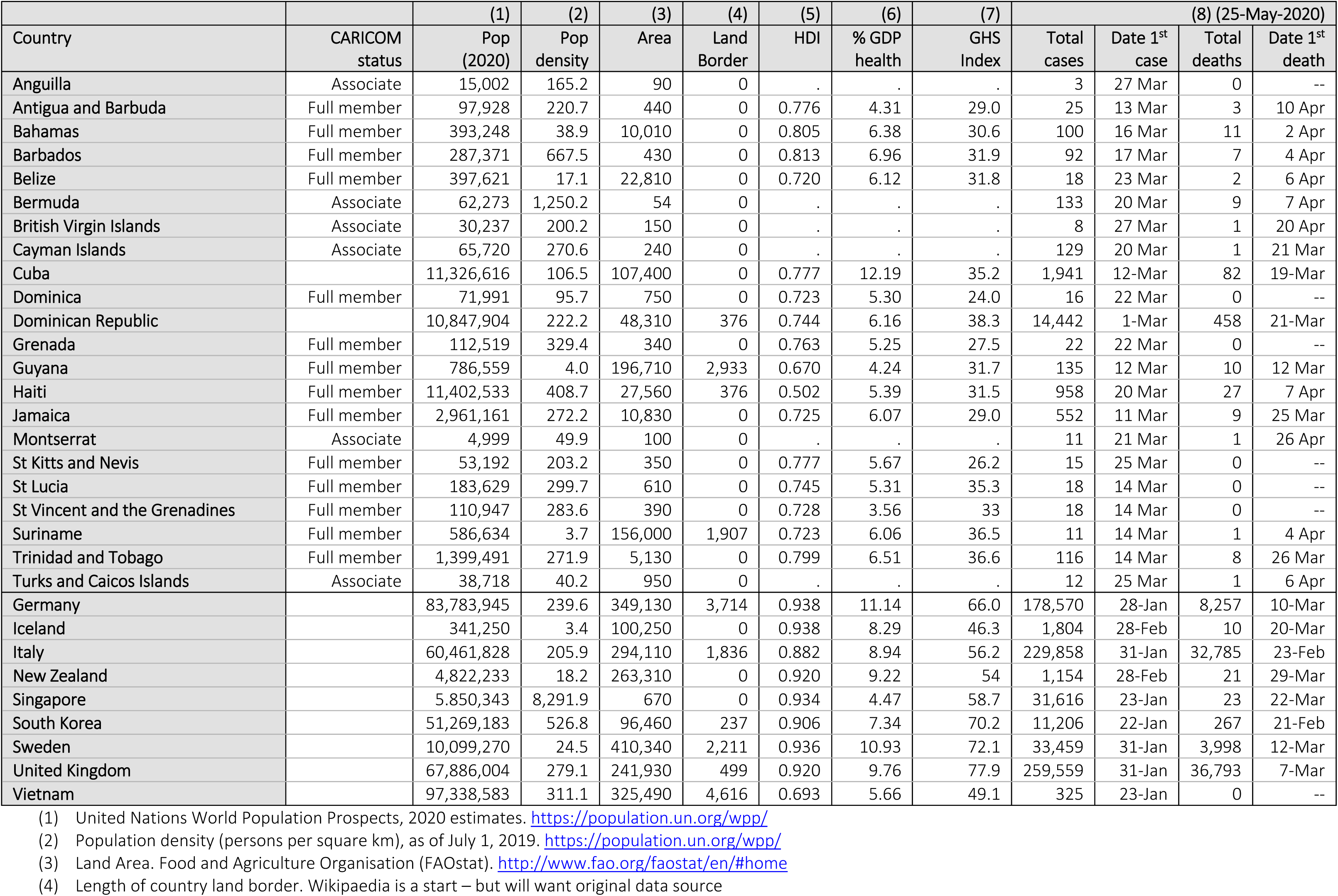
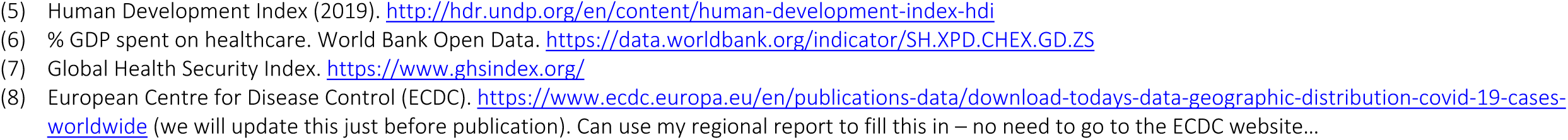
Selected characteristics of 22 Caribbean territories and 9 international locations

| Country | CARICOM status | (1)<br>Pop<br>(2020) | (2)<br>Pop<br>density | (3)<br>Area | (4)<br>Land<br>Border | (5)<br>HDI | (6)<br>% GDP<br>health | (7)<br>GHS<br>Index | (8) (25-May-2020) |  |  |  |
| --- | --- | --- | --- | --- | --- | --- | --- | --- | --- | --- | --- | --- |
|  |  |  |  |  |  |  |  |  | Total<br>cases | Date 1<sup>st</sup><br>case | Total<br>deaths | Date 1<sup>st</sup><br>death |
| Anguilla | Associate | 15,002 | 165.2 | 90 | 0 | . | . | . | 3 | 27 Mar | 0 | -- |
| Antigua and Barbuda | Full member | 97,928 | 220.7 | 440 | 0 | 0.776 | 4.31 | 29.0 | 25 | 13 Mar | 3 | 10 Apr |
| Bahamas | Full member | 393,248 | 38.9 | 10,010 | 0 | 0.805 | 6.38 | 30.6 | 100 | 16 Mar | 11 | 2 Apr |
| Barbados | Full member | 287,371 | 667.5 | 430 | 0 | 0.813 | 6.96 | 31.9 | 92 | 17 Mar | 7 | 4 Apr |
| Belize | Full member | 397,621 | 17.1 | 22,810 | 0 | 0.720 | 6.12 | 31.8 | 18 | 23 Mar | 2 | 6 Apr |
| Bermuda | Associate | 62,273 | 1,250.2 | 54 | 0 | . | . | . | 133 | 20 Mar | 9 | 7 Apr |
| British Virgin Islands | Associate | 30,237 | 200.2 | 150 | 0 | . | . | . | 8 | 27 Mar | 1 | 20 Apr |
| Cayman Islands | Associate | 65,720 | 270.6 | 240 | 0 | . | . | . | 129 | 20 Mar | 1 | 21 Mar |
| Cuba |  | 11,326,616 | 106.5 | 107,400 | 0 | 0.777 | 12.19 | 35.2 | 1,941 | 12-Mar | 82 | 19-Mar |
| Dominica | Full member | 71,991 | 95.7 | 750 | 0 | 0.723 | 5.30 | 24.0 | 16 | 22 Mar | 0 | -- |
| Dominican Republic |  | 10,847,904 | 222.2 | 48,310 | 376 | 0.744 | 6.16 | 38.3 | 14,442 | 1-Mar | 458 | 21-Mar |
| Grenada | Full member | 112,519 | 329.4 | 340 | 0 | 0.763 | 5.25 | 27.5 | 22 | 22 Mar | 0 | -- |
| Guyana | Full member | 786,559 | 4.0 | 196,710 | 2,933 | 0.670 | 4.24 | 31.7 | 135 | 12 Mar | 10 | 12 Mar |
| Haiti | Full member | 11,402,533 | 408.7 | 27,560 | 376 | 0.502 | 5.39 | 31.5 | 958 | 20 Mar | 27 | 7 Apr |
| Jamaica | Full member | 2,961,161 | 272.2 | 10,830 | 0 | 0.725 | 6.07 | 29.0 | 552 | 11 Mar | 9 | 25 Mar |
| Montserrat | Associate | 4,999 | 49.9 | 100 | 0 | . | . | . | 11 | 21 Mar | 1 | 26 Apr |
| St Kitts and Nevis | Full member | 53,192 | 203.2 | 350 | 0 | 0.777 | 5.67 | 26.2 | 15 | 25 Mar | 0 | -- |
| St Lucia | Full member | 183,629 | 299.7 | 610 | 0 | 0.745 | 5.31 | 35.3 | 18 | 14 Mar | 0 | -- |
| St Vincent and the Grenadines | Full member | 110,947 | 283.6 | 390 | 0 | 0.728 | 3.56 | 33 | 18 | 14 Mar | 0 | -- |
| Suriname | Full member | 586,634 | 3.7 | 156,000 | 1,907 | 0.723 | 6.06 | 36.5 | 11 | 14 Mar | 1 | 4 Apr |
| Trinidad and Tobago | Full member | 1,399,491 | 271.9 | 5,130 | 0 | 0.799 | 6.51 | 36.6 | 116 | 14 Mar | 8 | 26 Mar |
| Turks and Caicos Islands | Associate | 38,718 | 40.2 | 950 | 0 | . | . | . | 12 | 25 Mar | 1 | 6 Apr |
| Germany |  | 83,783,945 | 239.6 | 349,130 | 3,714 | 0.938 | 11.14 | 66.0 | 178,570 | 28-Jan | 8,257 | 10-Mar |
| Iceland |  | 341,250 | 3.4 | 100,250 | 0 | 0.938 | 8.29 | 46.3 | 1,804 | 28-Feb | 10 | 20-Mar |
| Italy |  | 60,461,828 | 205.9 | 294,110 | 1,836 | 0.882 | 8.94 | 56.2 | 229,858 | 31-Jan | 32,785 | 23-Feb |
| New Zealand |  | 4,822,233 | 18.2 | 263,310 | 0 | 0.920 | 9.22 | 54 | 1,154 | 28-Feb | 21 | 29-Mar |
| Singapore |  | 5,850,343 | 8,291.9 | 670 | 0 | 0.934 | 4.47 | 58.7 | 31,616 | 23-Jan | 23 | 22-Mar |
| South Korea |  | 51,269,183 | 526.8 | 96,460 | 237 | 0.906 | 7.34 | 70.2 | 11,206 | 22-Jan | 267 | 21-Feb |
| Sweden |  | 10,099,270 | 24.5 | 410,340 | 2,211 | 0.936 | 10.93 | 72.1 | 33,459 | 31-Jan | 3,998 | 12-Mar |
| United Kingdom |  | 67,886,004 | 279.1 | 241,930 | 499 | 0.920 | 9.76 | 77.9 | 259,559 | 31-Jan | 36,793 | 7-Mar |
| Vietnam |  | 97,338,583 | 311.1 | 325,490 | 4,616 | 0.693 | 5.66 | 49.1 | 325 | 23-Jan | 0 | -- |

- (1) United Nations World Population Prospects, 2020 estimates. https://population.un.org/wpp/ - (2) Population density (persons per square km), as of July 1, 2019. https://population.un.org/wpp/ - (3) Land Area. Food and Agriculture Organisation (FAOstat). http://www.fao.org/faostat/en/#home - (4) Length of country land border. Wikipaedia is a start – but will want original data source
- (5) Human Development Index (2019). http://hdr.undp.org/en/content/human-development-index-hdi - (6) % GDP spent on healthcare. World Bank Open Data. https://data.worldbank.org/indicator/SH.XPD.CHEX.GD.ZS - (7) Global Health Security Index. https://www.ghsindex.org/ - (8) European Centre for Disease Control (ECDC). https://www.ecdc.europa.eu/en/publications-data/download-todays-data-geographic-distribution-covid-19-cases-worldwide (we will update this just before publication). Can use my regional report to fill this in – no need to go to the ECDC website...

In Figure 1 we visualise the cumulative numbers of confirmed cases and deaths across the Caribbean. As of Monday 25th May, there were 18,755 confirmed cases and 631 confirmed deaths among the 22 Caribbean countries and territories. Confirmed cases were dominated by Dominican Republic (14,422 cases, 76.9% of all cases) and to a lesser extent by Cuba (1,941 cases, 10.3% of all cases). The remaining 20 CARICOM countries and territories accounted for 2,392 confirmed cases, or 12.8% of all cases. Similarly, confirmed deaths were dominated by Dominican Republic (458 deaths, 72.6% of all deaths) and to a lesser extent by Cuba (82 deaths, 13.0% of all deaths). The 20 CARICOM countries and territories accounted for 91 confirmed deaths, or 14.4% of all deaths.

**Figure 1.**
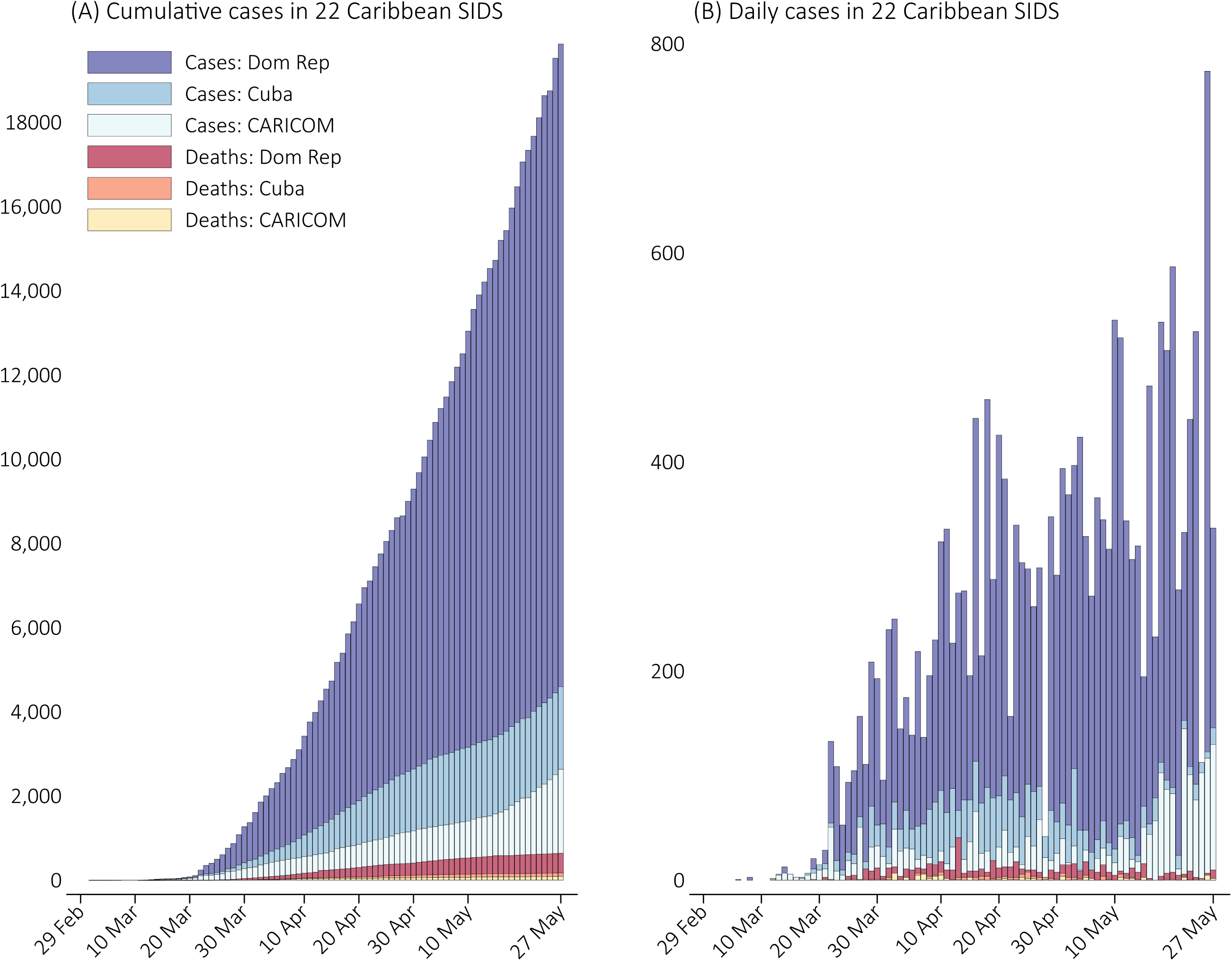
Numbers of confirmed cases and confirmed deaths from COVID-19 in 22 Caribbean countries and territories up to 25 May 2020

In Figure 2 we visualise outbreak growth rates by country. Growth rates varied markedly over time in most countries and territories, reflecting periods in each country when higher or lower numbers of cases were identified. CARICOM countries experienced the outbreak later than comparator countries and have so far maintained lower levels of growth than those seen in comparator countries. As of 25-May-2020, twelve out of 22 Caribbean territories had kept their maximum growth rates below 10% and of the remaining ten Caribbean territories, maximum growth rates ranged between 13% (The Bahamas, Jamaica) and 40% (Dominican Republic). Most Caribbean growth rate trajectories were similar in magnitude to those seen in two Asian comparator countries, Vietnam (13%) and Singapore (16%), indicative of good initial outbreak control. Comparator countries saw higher growth rates and a wider range of growth, between 13% (Vietnam) and 63% (Italy).

**Figure 2.**
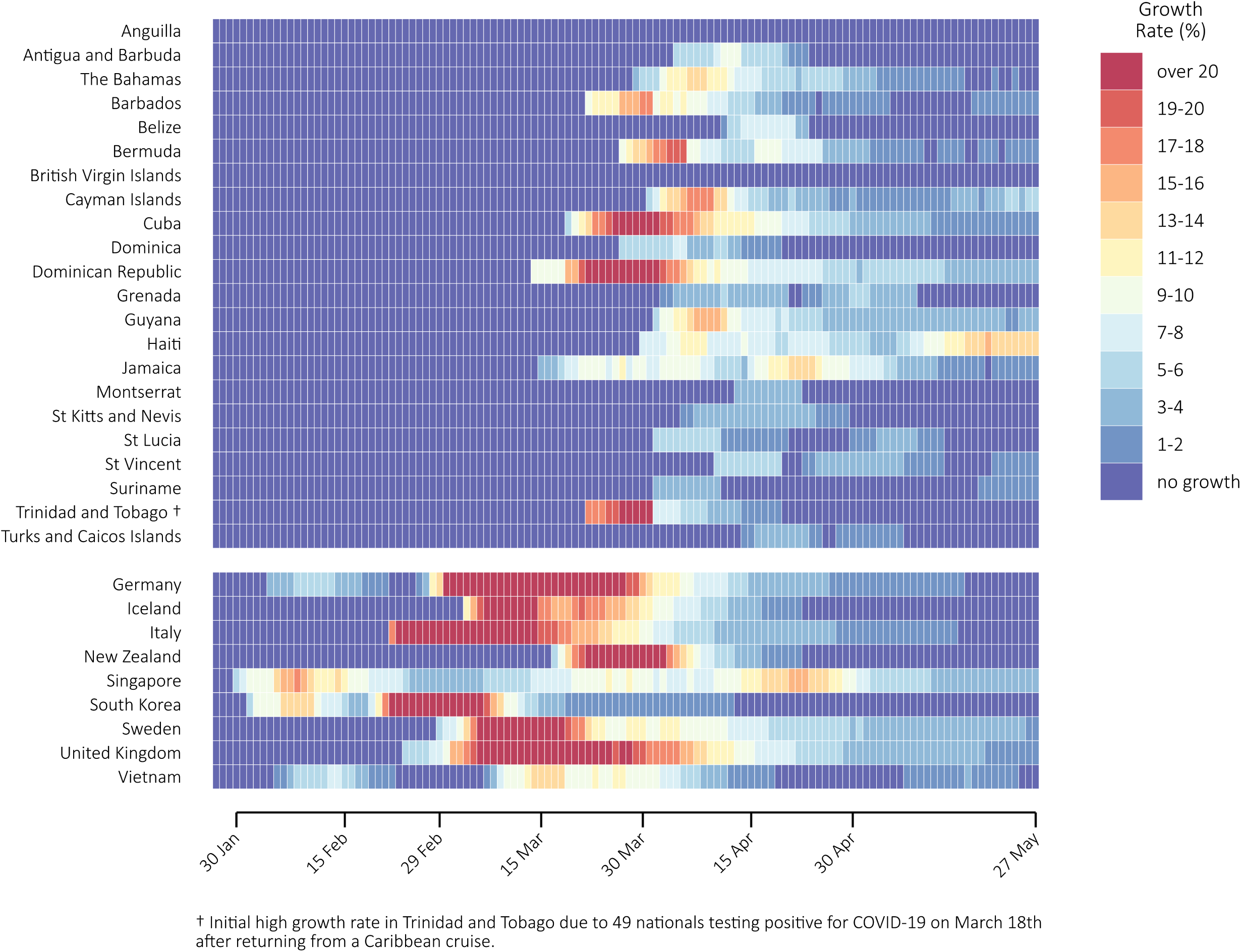
COVID-19 confirmed case growth rates in 22 Caribbean countries and territories and 9 international locations up to 25-May-2020

In Figure 3 we present containment measures in our three broad categories: measures to control movement into a country, measures to control movement within a country, and measures to control mass gatherings. Sixteen out of the 22 Caribbean countries implemented a full border closure, compared to 1 of 9 comparator countries (New Zealand). Roughly equal proportions of Caribbean and comparator countries initiated some form of lockdown, but only Caribbean countries implemented strict night-time curfews. All countries implemented measures to control gatherings.

**Figure 3.**
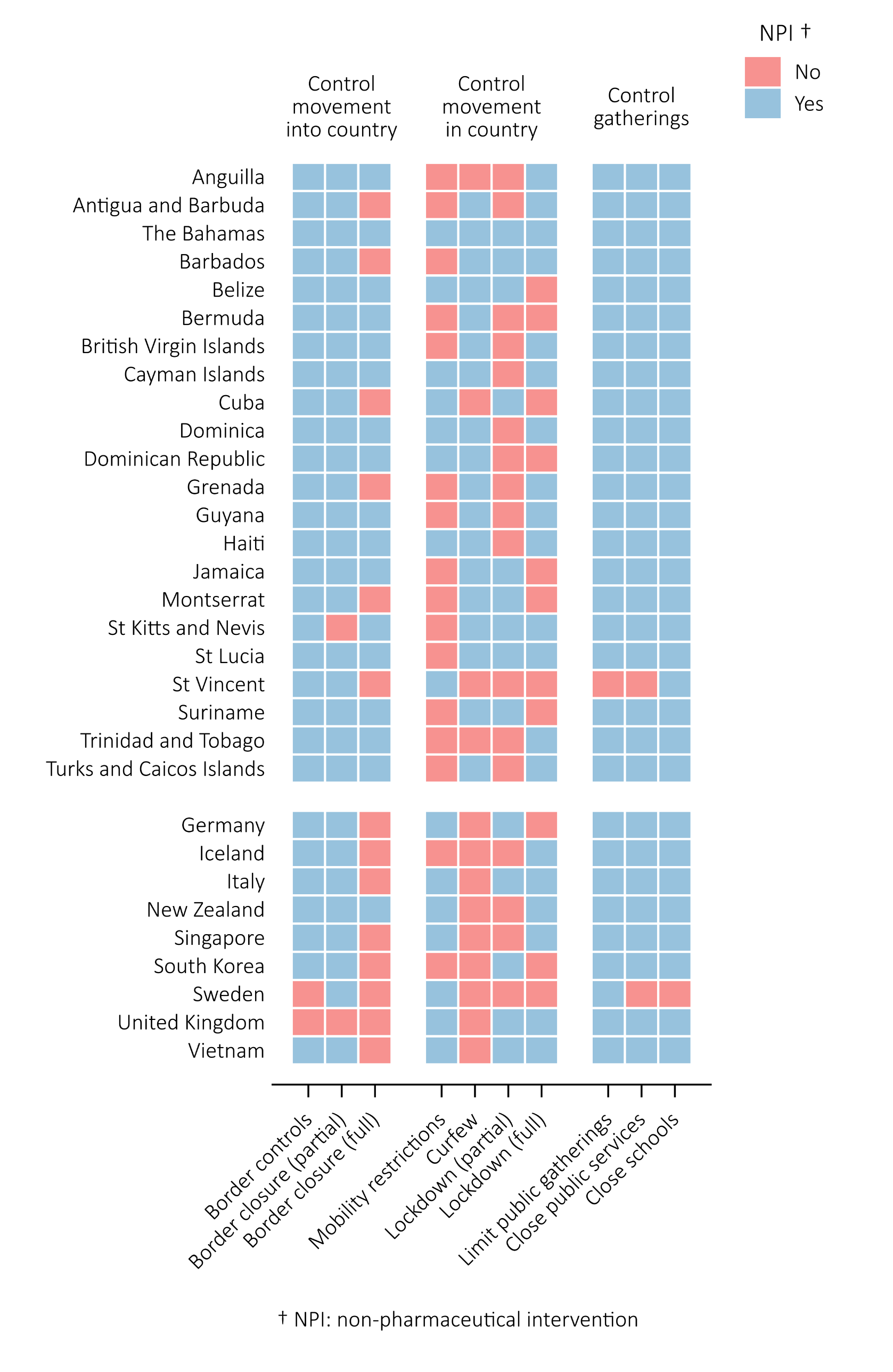
Non pharmaceutical interventions (NPIs) implemented by 22 Caribbean countries and territories and 9 international locations, stratified by type of NPI.

In Figure 4 we present the timing of NPIs, relative to the date of first confirmed case in each country. Broadly, Caribbean countries and territories tended to implement NPIs earlier, compared to the international comparator countries. Within the Caribbean the order of implementing measures has been control of movement into countries, followed by control of gatherings, and then control of movement within countries. Comparator countries tended to follow the same pattern but waited longer before implementation. When examining the spread of NPI timings across the Caribbean, many Caribbean territories followed similar timings to New Zealand and Iceland. On average, Caribbean countries began controlling movements into countries 27 days before the first confirmed case (inter-quartile range (IQR) 48 to 4 days before). This compares to 4 days before the first confirmed case among comparator countries (IQR 23 days before to 22 days after). Caribbean countries began controlling movement within a country 9 days after the first confirmed case (IQR 2 days to 15 days after), compared to 45 days after among comparator countries (IQR 24 to 55 days after). Caribbean countries began controlling gatherings 1 day before the first confirmed case (IQR 5 days before to 2 days after), compared to 29 days after among comparator countries (IQR 22 to 42 days after).

**Figure 4.**
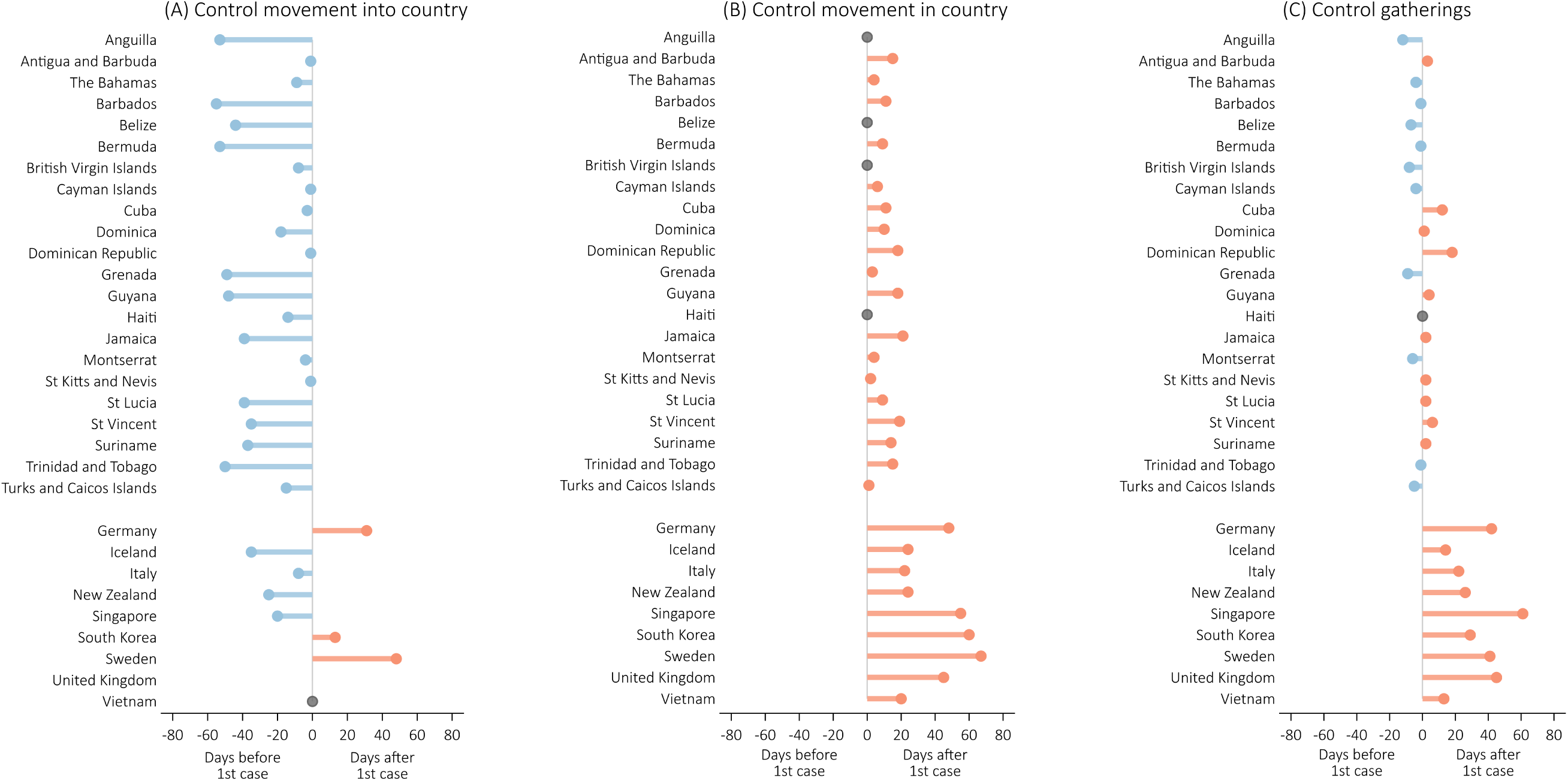
The number of days between the date of first confirmed case of COVID-19 and the date of introducing the first non-pharmaceutical intervention (NPI), stratified by country and by NPI type.

In Figure 5 we present the maximum reduction in weekly community movement data in selected Caribbean and comparator countries. With the exception of Haiti (39% maximum reduction) and Jamaica (50% maximum reduction), the largest weekly reductions in movement over a full week were above 60% in all Caribbean countries, over 70% in 5/8 Caribbean countries, and over 80% in 1 Caribbean country (Barbados). In the 8 comparator countries with available movement data, maximum movement reductions were over 70% in only 2/8 countries (Italy, New Zealand). The implementation of a major NPI to limit movement (curfews and/or lockdowns) was largely followed by a fall in population mobility, but there was much variation in this effect. Several countries saw a sharp fall in mobility co-incident with the date of either curfew and/or lockdown implementation (Antigua and Barbuda, Barbados, Trinidad and Tobago, New Zealand, Singapore), while for others the decline was either more gradual or less pronounced (see Supplement graphics for country-level details). In Barbados (for example) two drops in movement occurred: a 23 percentage point drop on 29^th^ March, 1-day after a national curfew order, and a further 32 percentage point drop on 4^th^ April, 1 day after a national stay-at-home order. Similarly, stark drops were seen in Antigua and Barbuda, New Zealand and Italy, all of which implemented stringent lockdown conditions (see Supplement for details).

**Figure 5.**
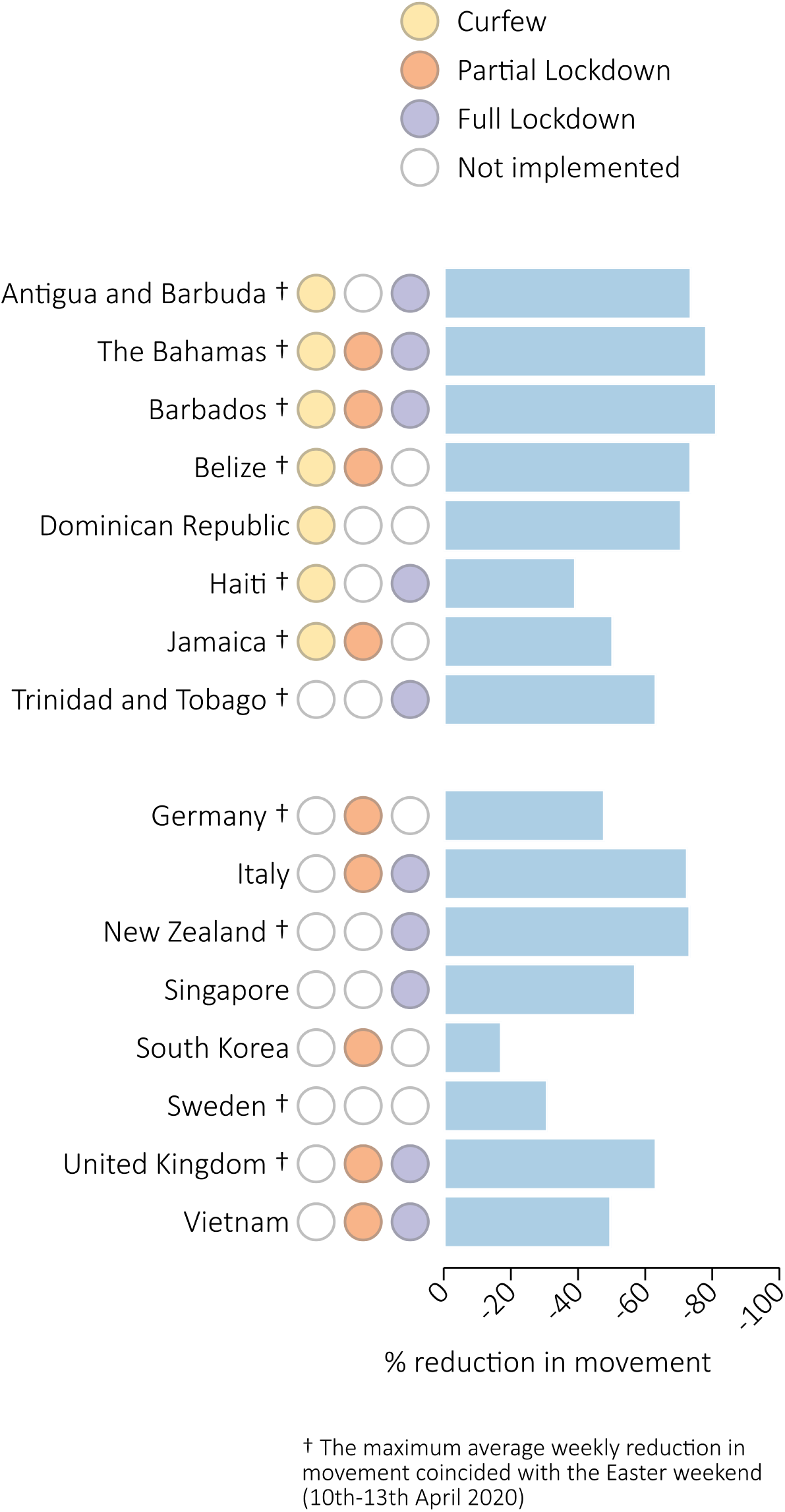
The maximum average weekly reduction in population mobility among 8 Caribbean countries and 8 comparator countries.

## DISCUSSION

As of Monday 25th May, there were 18,755 confirmed cases and 631 confirmed deaths among the 22 Caribbean countries and territories. The maximum growth rate in the Caribbean was 40% in the Dominican Republic, compared to 63% in the comparator country of Italy. Caribbean countries were more likely to implement a full border closure and strict night-time curfews. They were also more likely to introduce NPI measures earlier than comparator countries relative to the date of their first confirmed case. Peak movement reductions were above 70% in 5/8 Caribbean countries, and in only 2/8 comparator countries.

As of 25 May 2020, the Caribbean region has broadly achieved initial COVID-19 containment, with the exception of the evolving outbreak on the island of Hispaniola. Given the struggles some developed comparator countries have had with containment, the low GHS scores across the Caribbean, and the benefits of avoiding community transmission due to high national population densities, this is a significant success for the CARICOM SIDS. Although Caribbean countries initially developed their NPI implementation strategies using local expertise and international evidence, they nevertheless followed similar initial NPI pathways, focusing first on measures to limit movement into their respective countries. These sensible early precautions were an attempt to block imported cases and may be particularly important in small islands with a limited and manageable number of physical entry points. The significance of early border controls and closures should be placed within the economic context of the region. Tourism is a dominant revenue stream for many Caribbean SIDS, with their reliance on international arrivals, particularly from Europe and North America. Governments were aware that border controls and closures would have severe economic effects. Weighed against this was the known fragility of regional health systems, and governments were keen to avoid their health systems being overwhelmed by a sharp increase in hospitalisations. Using the date of first confirmed case in each country as our indicator, Caribbean SIDS generally implemented NPIs earlier than our chosen comparator countries. For movements into a country, the Caribbean on average implemented controls 23 days before their comparator counterparts. For control of movement within countries, the Caribbean implemented controls 36 days before comparators, and for control of gatherings the Caribbean on average implemented controls 30 days before comparator countries. This three-to-four week “head-start” by Caribbean countries may be partly attributed to the region having seen the outbreak unfold in other parts of the world, and the longer ‘grace period’ before COVID-19 arrival in the region. It may also reflect Governments’ recognising the potential for hospitalisations to overwhelm vulnerable health system infrastructures, spurring them into strong and early outbreak suppression. (19)

Of the 193 countries in the ACAPS database, 96 initiated a curfew order, with 59% of those countries in Africa and the Americas. In the Caribbean, governments quickly passed emergency laws that allowed for enforceable curfews and stay-at-home orders. Curfews were common in the Caribbean, with populations not allowed to leave their homes for any reason except emergencies (or emergency work). Associated punitive measures were regularly significant; in Barbados for example, a fine of 50,000 Barbados dollars (USD 25,000) or 1-year in prison were possible. Curfews were mostly applied during the hours of darkness, and a logic to this would have been be an attempt to prevent evening gatherings. Although curfews were not officially implemented in any of our comparator countries, this may reflect the semantics of terminology, with curfews possibly seen by some governments as sounding overly authoritarian. Some countries, without using the term “curfew”, operated near curfew-like conditions. Italy for example required those leaving their homes to carry movement exemption forms, with fines for breaches of these rules. The stringency of national lockdowns, including curfews seem to impact heavily on community mobility, with countries implementing strict measures seeing stark drops in post-implementation mobility. The extent of a Government’s willingness to implement and enforce stringent movement restrictions will have been a compromise between the desire to limit transmission and the perceived success of the intervention given known societal norms. A full examination of these influences is important, but beyond the scope of this initial work. Among countries for which human mobility data were available (8 Caribbean countries, 8 comparators) curfews and lockdowns were visually associated with marked falls in human movement. For example, Barbados, Antigua and Barbuda, Trinidad and Tobago, and New Zealand saw sustained post-lockdown drops in excess of 30 percentage points (see supplement for details). In the case of Barbados, implementation of curfew followed by lockdown each coincided with clear mobility reductions. It may be that a curfew order acted as a national sensitisation measure for subsequent full lockdown.

For the Caribbean, the coming weeks and months are likely to involve discussions and actions centred around easing national NPIs while maintaining broad outbreak containment. A priority is how to safely but effectively re-invigorate international tourism as Caribbean islands look to reopen their economies for business. Increased tourism from the European and North American markets, which are still grappling to contain the virus, increases the opportunity for imported cases and local transmission. Consequently, modified NPIs that minimise the chance of a renewed outbreak without negatively impacting the tourist experience will need to be envisaged. One potential option is the concept of travel corridors allowing free movement between countries or cities that have good containment, but restricting movement from higher-risk locations to safeguard public health. As timing of NPIs has emerged as an important factor in containing the outbreak, it should now encourage a proactive approach as countries plan to encourage tourism. National evidence-based risk assessments, drawing on country-level goals and limitations must now be a priority. Continued outbreak surveillance remains a critical tool to enable swift action following accelerated transmission. As always, it could be sensible to learn from successful models implemented elsewhere.

This descriptive study has limitations. We focus only on NPIs related to movement and we recognize that other NPIs such as contact tracing, and isolation and quarantine protocols will have also helped to reduce transmission. We have described the type and timing of NPIs implemented and have inferred their impact on COVID-19 transmission. However, we have not attempted to quantify these effects, nor have we accounted for other factors that may impact transmission, such as the effect of climate on virus longevity. (20) The data we use in this article has been drawn from disparate sources, each of which has limitations. The number of cases we report is based on the number of tests performed, and as population-level testing remains economically impractical, no country currently knows the extent of their underlying outbreak. The NPI data are drawn partly from informal sources such as media reports with implied quality concerns. To counter this we have made systematic efforts to triangulate our NPI information wherever possible. Although we have used two quantitative categories to describe the implementation of lockdowns, in reality national stay-at-home orders varied markedly in their stringency and geographical reach. This variation in stringency included country-level differences in what constituted essential services.

Stringent controls to limit movement, and specifically the early timing of those controls has had an important impact on containing the spread of COVID-19 across much of the Caribbean. Very early controls to limit movement into the country may well be particularly effective for SIDS. With much of the region economically reliant on international tourism, and with steps to open borders now being considered, it is critical that the region draws on a solid evidence-base to balance the competing demands of economics and public health.

## Data Availability

This secondary analysis involves the use of four sources of open access data. They are available at the following online locations. The sources are as follows: SOURCE 1. Non-pharmceutical interventions ACAPS COVID-19 Government Measures Database. [Available from: https://www.acaps.org/covid19-government-measures-dataset. SOURCE 2. COVID-19 cases and deaths European Centre for Disease Control (ECDC). Daily data download on the geographic distribution of COVID19 cases worldwide. [Available from: https://www.ecdc.europa.eu/en/publications-data/download-todays-data-geographic-distribution-covid-19-cases-worldwide. SOURCE 3. COVID-19 cases and deaths Johns Hopkins coronavirus Resource Center. [Available from: https://coronavirus.jhu.edu/map.html. SOURCE 4. Community mobility Google LLC. COVID-19 Community Mobility Reports [Available from: https://www.google.com/covid19/mobility/index.html?hl=en. We supplemented the ACAPS dataset with new information from 6 United Kingdom Overseas Territories. Our full dataset for 31 included countries and 10 non-pharmaceutical interventions (310 data rows) is available at the following temporary location: https://moodle.caribdata.org/lms/enrol/index.php?id=27

---

**Supplement 1.**
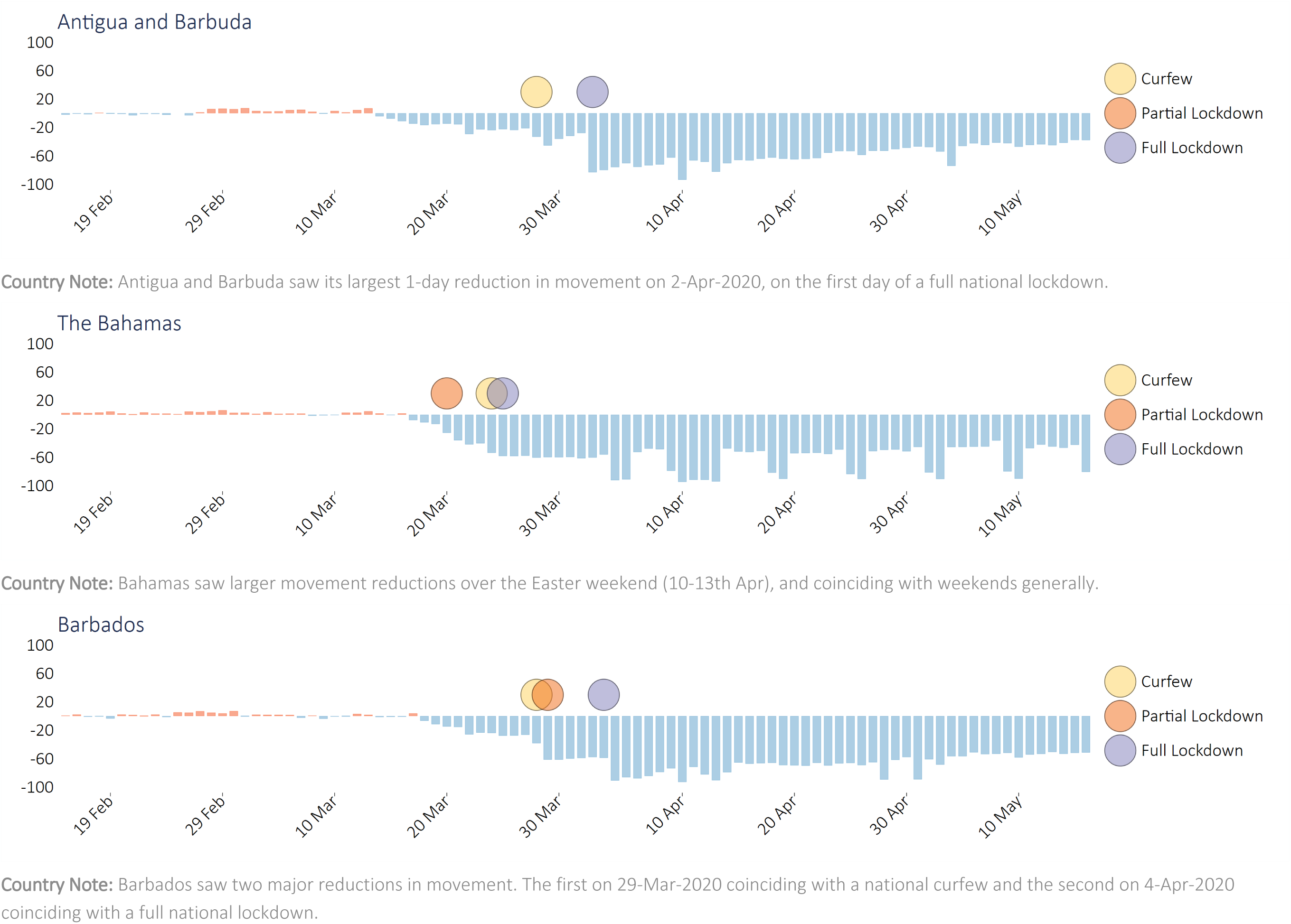
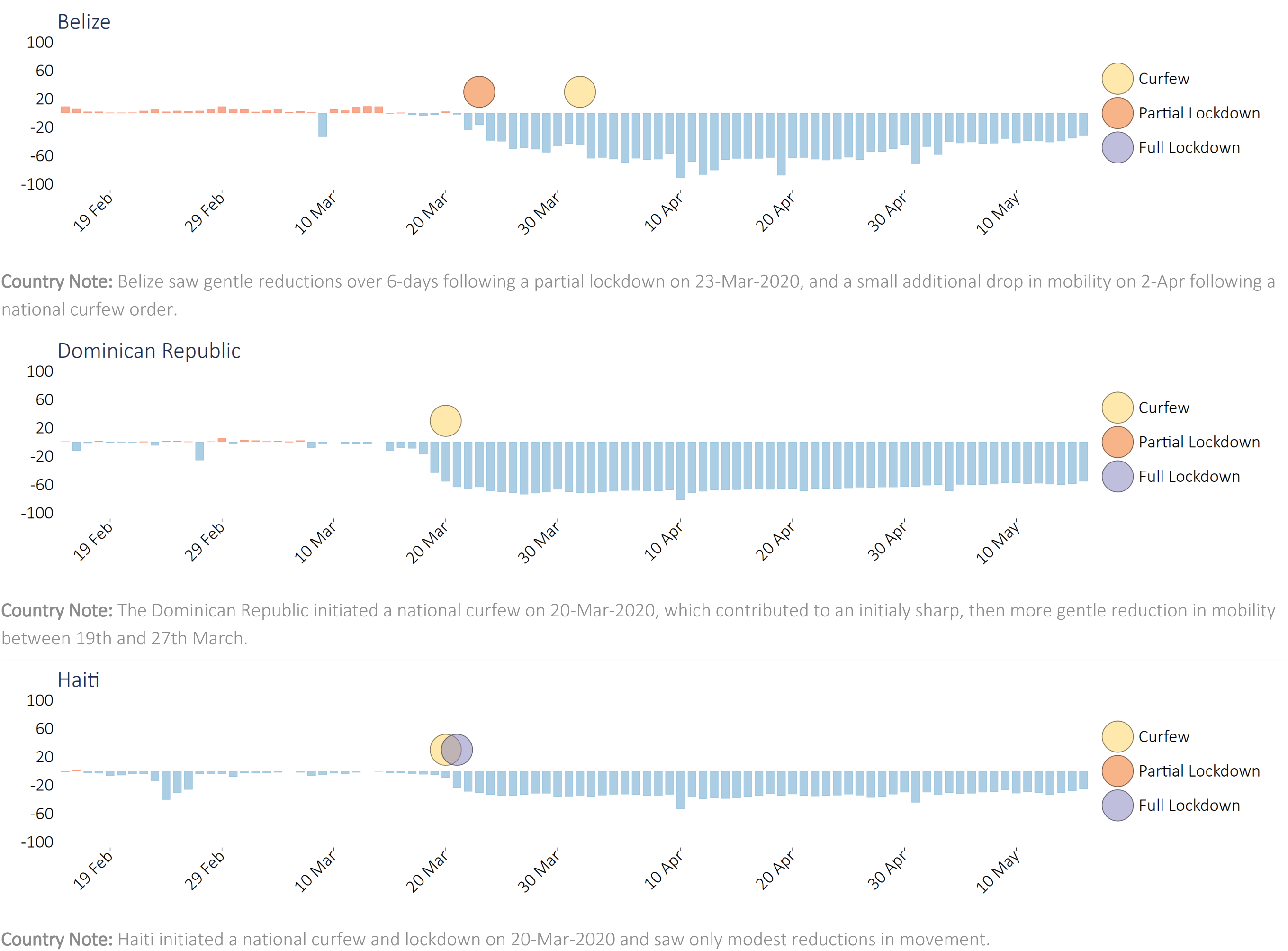
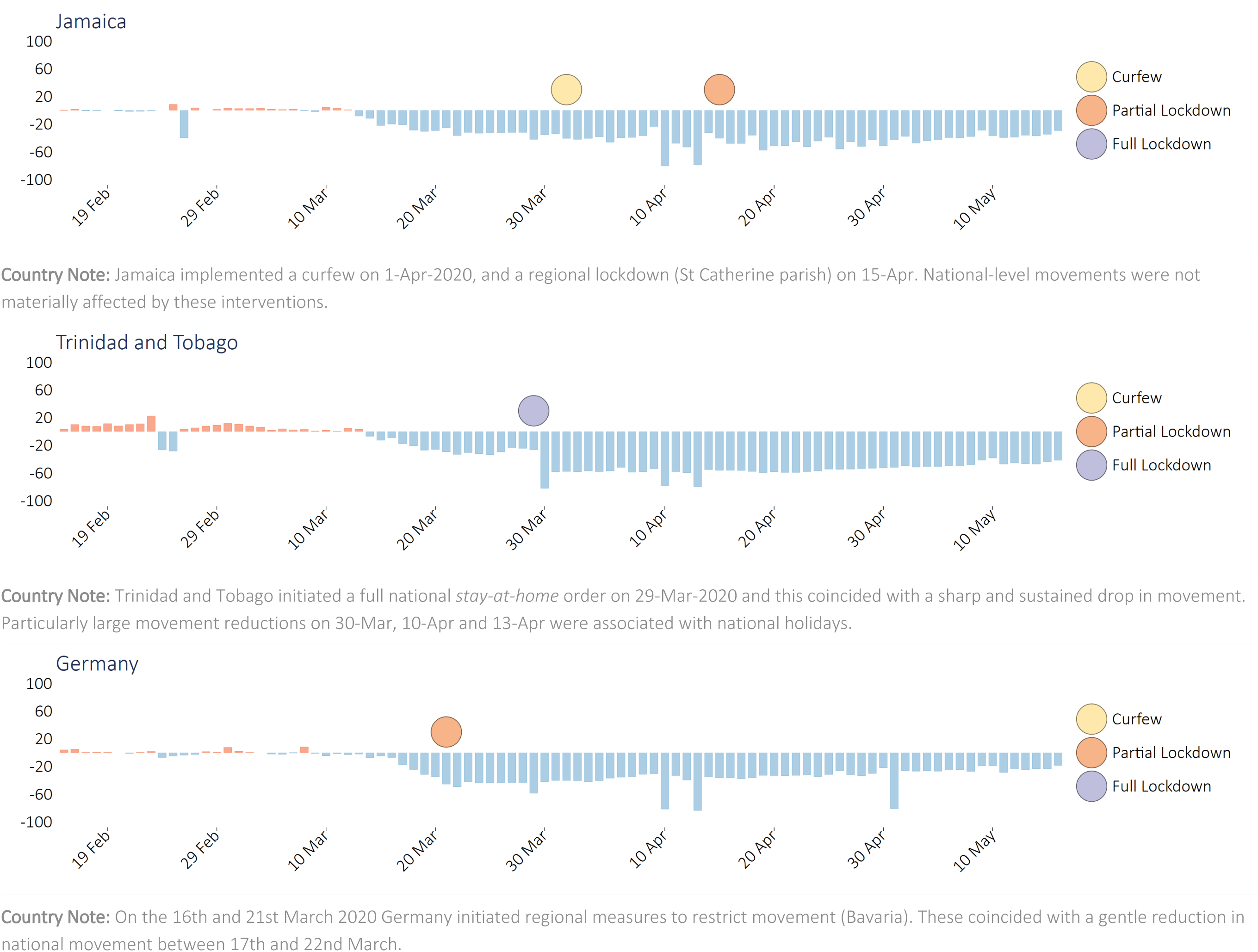
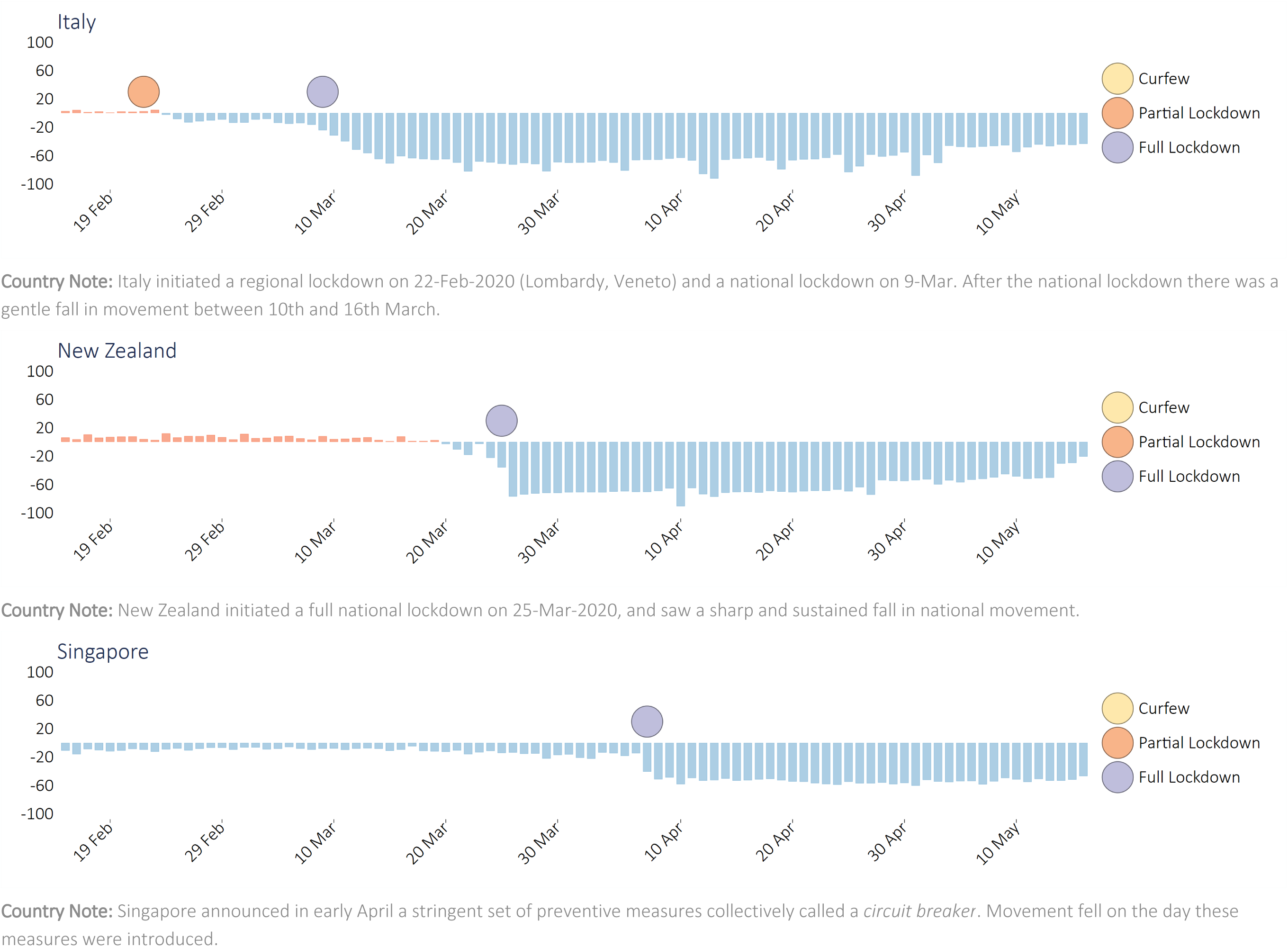
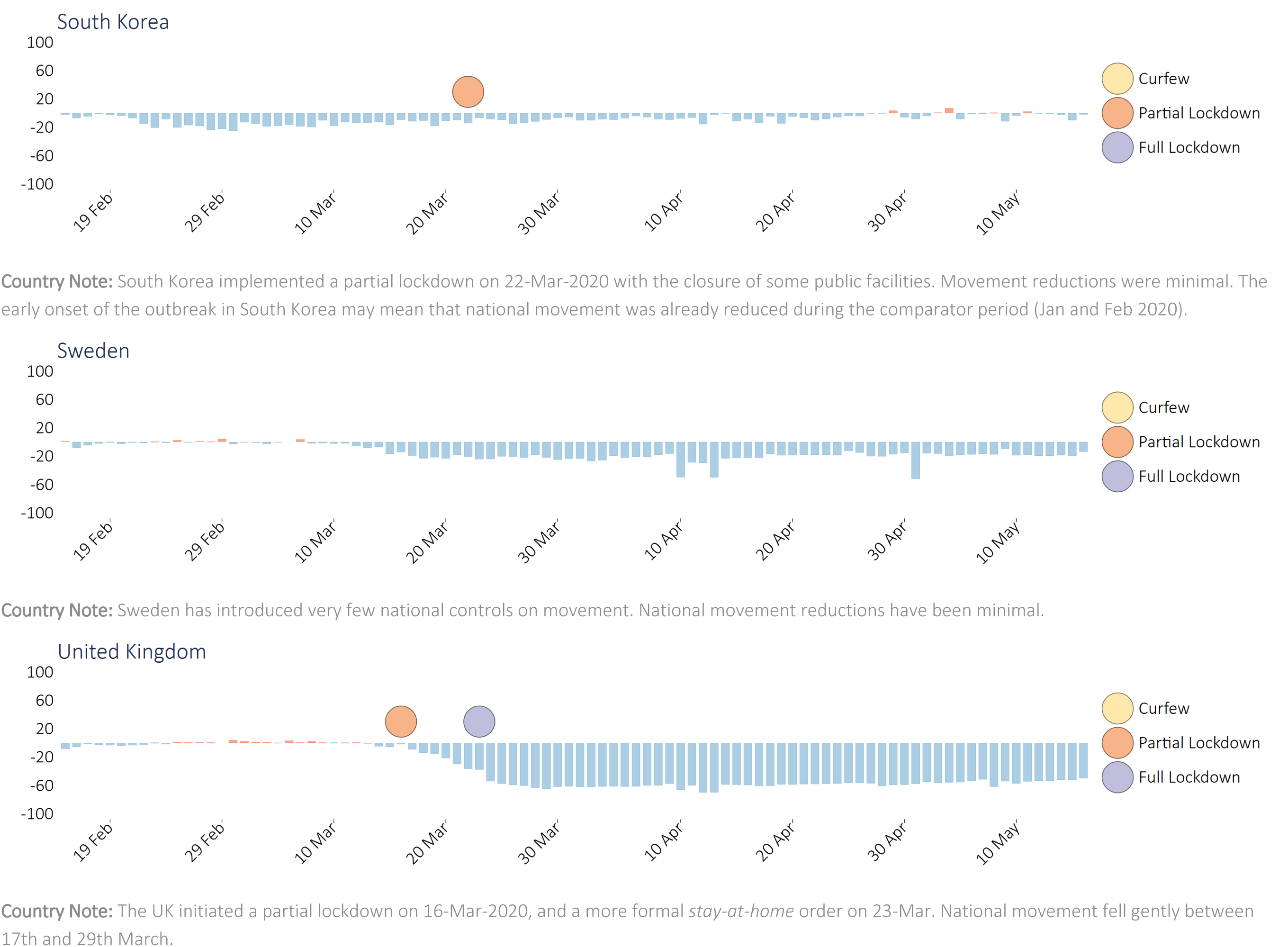
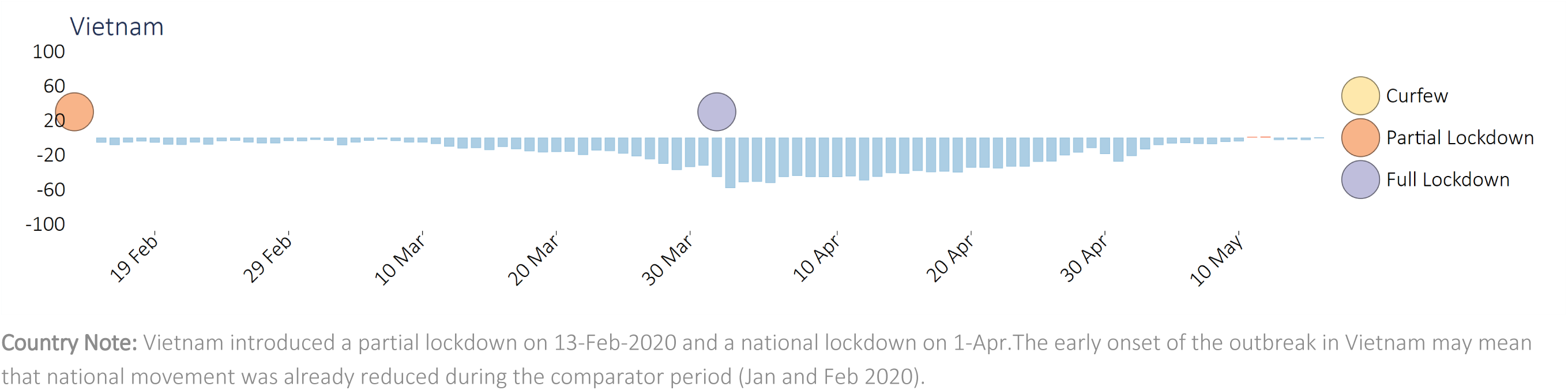
Changes in community movement and dates of national curfews and lockdowns among 8 Caribbean countries and 8 comparator countries.

**Supplement 2**. Protocol describing systematic search for non‐pharmaceutical interventions in 22 Caribbean countries and 9 international comparator countries.

## Background

The ACAPS COVID-19 government measures database^1^ collates non-pharmaceutical interventions (NPIs) implemented by governments worldwide in response to the coronavirus epidemic. It uses a variety of international and national media sources and includes a secondary review of collated information. Measures are grouped into 5 broad categories: movement restriction, social distancing, public health measures, governance and socioeconomic measures, and lockdown. Given the rapidly changing nature of national interventions – especially early in the crisis – and the use of unverified data sources, we aimed to validate ACAPS for the Caribbean region and our 9 chosen comparator countries by conducting a review of all listed non-pharmaceutical interventions (NPIs) and systematically search for missing entries.

## Methods

### Eligibility criteria

#### Countries included

*Caribbean:* Anguilla, Antigua and Barbuda, Bahamas, Barbados, Belize, Bermuda, British Virgin Islands, Cayman Islands, Cuba, Dominica, Dominican Republic, Grenada, Guyana, Jamaica, Haiti, Montserrat, Saint Kitts and Nevis, Saint Lucia, Saint Vincent and the Grenadines, Suriname, Trinidad and Tobago, and Turks and Caicos

*Comparator:* Germany, Iceland, Italy, New Zealand, Singapore, South Korea, Sweden, United Kingdom, Vietnam

#### Definitions of measures

Of the 34 NPI groupings detailed in the ACAPS database, 15 are related to limiting human movement into a country, limiting movement within a country, or limiting gatherings (Table 1, Column 1). We collapsed these 15 ACAPS measures into the 10 NPI sub-categories (Column 2), and 3 NPI major categories (Column 3). Our 10 NPI sub-categories are defined in full in Column 4, and we used these definitions when categorizing individual national NPI measures (see Table 1).

**Table 1:** Composition of 15 original ACAPS NPI categories, 3 major NPI categories, and 10 NPI sub-categories, along with associated definitions

| Original ACAPS categories used<br>(15 applicable categories) | 10 NPI Sub-Categories | 3 NPI Major Category | Definition of each NPI sub-category |
| --- | --- | --- | --- |
| Additional health documents required on arrival<br>Health screenings in airports<br>Border checks<br>Visa restrictions | Border Control | **Movement into country** | Any measure to intensify border controls, including health checks at border, visa restrictions or suspensions, and requirements for additional health documents. |
| International flight suspensions<br>Border closures | Border Closure: Full |  | All points of entry closed including complete suspension of international flights and/or full airport closure |
| International flight suspensions<br>Border closures<br>Complete border closure | Border Closure: Partial |  | Some points of entry into country closed and/or passengers from certain destinations not permitted entry. This also includes any flight suspensions from specific destinations |
| Domestic travel restriction<br>Checkpoints within country | Mobility restrictions | **Movement within country** | Includes domestic travel restrictions with or without structured or ad-hoc security checks. |
| Curfew<br>Partial lockdown | Curfews |  | Government order for people to remain in their home between specified hours (mostly at night). |
| Curfew<br>Partial lockdown | Lockdown: full |  | 24-hour curfew and/or country under emergency “*stay at home*” order, and closure of public spaces. Only movement of essential workers allowed. Only essential services open. |
| Curfew<br>Partial lockdown<br>Full lockdown | Lockdown: partial |  | As full lockdown, except that some public spaces remain open and/or specific businesses remain open, in addition to essential services. |
| Limit public gatherings | Limit public gatherings | **Limit public gatherings** | Any measure to ban or reduce the number of people allowed at public gatherings (such as weddings, funerals, religious worship) and social occasions. |
| Closure of businesses and public services | Closure of businesses & public services |  | Any measure to close or limit public access to non-healthcare public services, and/or private businesses. |
| Schools closure | Close schools |  | Any measure to close an educational facility, including tertiary education |

##### Search strategy, data extraction

We constructed a new dataset comprising 1 row of information for each country / NPI sub category combination (31 countries and 10 NPI sub-categories, creating a dataset of 310 rows of information). Using the ACAPS database as the major information source, and supplementing this source with a predefined list of additional sources (see Table 2), for each country we documented the existence of NPI measures in each NPI sub-category, recording the date of implementation, the description of the NPI measure, and the data source. This search strategy was performed by two reviewers (MM, KR), with a third reviewer (CH) adjudicating on all discrepant information.

##### Synthesis of results

This review was planned as a narrative synthesis of evidence, summarizing the NPIs implemented in each country by the major NPI categories and NPI sub-categories described in Table 1. Further information on data analytics associated with the collected NPI data is provided in the main article (Methods / Statistical Methods).

**Table 2:** Data sources used during systematic literature search for national non-pharmaceutical interventions

| COUNTRY | GOVERNMENT | TOURIST INFORMATION AND EMBASSIES | NATIONAL NEWSPAPERS AND WIKIPEDIA |
| --- | --- | --- | --- |
| Anguilla | http://www.gov.ai/covid.php | https://www.gov.uk/foreign-travel-advice/anguilla<br>https://bb.usembassy.gov/u-s-citizen-services/covid-19-information | www.thedailyherald.sx |
| Antigua and Barbuda | https://ab.gov.ag | https://www.gov.uk/foreign-travel-advice/antigua-and-barbuda<br>https://bb.usembassy.gov/u-s-citizen-services/covid-19-information | https://antiguaobserver.com |
| Bahamas | www.bahamas.gov.bs | https://bs.usembassy.gov/covid-19-information/<br>https://www.gov.uk/foreign-travel-advice/bahamas | https://en.wikipedia.org/wiki/COVID-19_pandemic_in_the_Bahamas<br>http://www.tribune242.com |
| Barbados | https://gisbarbados.gov.bb | https://bb.usembassy.gov/u-s-citizen-services/covid-19-information/ | https://barbadostoday.bb<br>https://www.barbadosadvocate.com<br>https://www.nationnews.com |
| Belize | https://www.pressoffice.gov.bz<br>http://health.gov.bz/www/ | https://belizetourismboard.org/belize-covid-19-update-for-travellers/<br>http://belizehighcommission.co.uk/ | http://belizenews.com<br>https://amandala.com.bz/news |
| Bermuda | https://www.gov.bm/coronavirus | https://www.gotobermuda.com/bta<br>https://bm.usconsulate.gov/u-s-citizen-services/security-and-travel-information | http://www.royalgazette.com |
| British Virgin Islands | https://bvi.org.uk | https://bb.usembassy.gov/u-s-citizen-services/covid-19-information<br>http://www.bvi.gov.vg/departments/bvi-tourist-board | http://www.virginislandsnews.com |
| Cayman Islands | http://www.gov.ky/portal/page/portal/cighome/pressroom | https://jm.usembassy.gov/covid-19-information-cayman-islands/<br>https://www.explore.gov.ky | https://www.caymaniantimes.ky<br>https://www.conyers.com |
| Cuba |  | https://www.gov.uk/foreign-travel-advice/cuba/coronavirus<br>https://cu.usembassy.gov/covid-19-information/ | http://www.cubadebate.cu<br>https://en.wikipedia.org/wiki/COVID-19_pandemic_in_Cuba |
| Dominica | http://dominica.gov.dm/corona | https://travel.state.gov/content/travel/en/international-travel/International-Travel-Country-Information-Pages/Dominica.html<br>https://www.gov.uk/foreign-travel-advice/dominica | https://dominicanewsonline.com |
| Dominican Republic | https://coronavirusrd.gob.do | https://www.gov.uk/foreign-travel-advice/dominican-republic/coronavirus<br>https://do.usembassy.gov/covid-19-information/ |  |
| Grenada | https://www.mgovernance.net | https://www.gov.uk/foreign-travel-advice/grenada | https://www.nowgrenada.com |
| Guyana | https://dpi.gov.gy | https://travel.state.gov/content/travel/en/traveladvisories/traveladvisories/guyana-travel-advisory.html | http://guyanachronicle.com/<br>https://www.stabroeknews.com/<br>https://www.kaieteurnews.com |
| Haiti |  | https://www.gov.uk/foreign-travel-advice/haiti/health<br>https://www.humanitarianresponse.info/en/operations/ha%C3%Afti | https://www.haitilibre.com<br>https://en.wikipedia.org/wiki/COVID-19_pandemic_in_Haiti |
| Jamaica | https://www.moh.gov.jm | https://jm.usembassy.gov/covid-19-information-jamaica<br>https://www.gov.uk/foreign-travel-advice/jamaica | http://jamaica-gleaner.com |
| Montserrat | http://www.gov.ms | https://www.gov.uk/foreign-travel-advice/montserrat<br>https://bb.usembassy.gov/u-s-citizen-services/covid-19-information<br>https://travel.state.gov/content/travel/en/international-travel/International-Travel-Country-Information-Pages/Montserrat.html | https://www.themontserratreporter.com |
| St Kitts and Nevis |  | https://www.gov.uk/foreign-travel-advice/st-kitts-and-nevis<br>https://bb.usembassy.gov/u-s-citizen-services/covid-19-information | https://www.thestkittsnevisobserver.com<br>https://en.wikipedia.org/wiki/COVID-19_pandemic_in_Saint_Kitts_and_Nevis<br>https://zizonline.com |
| St Lucia | www.govt.lc | https://www.gov.uk/foreign-travel-advice/st-lucia<br>https://bb.usembassy.gov/u-s-citizen-services/covid-19-information | https://www.loopslu.com |
| St Vincent and the Grenadines | http://health.gov.vc | https://bb.usembassy.gov/u-s-citizen-services/covid-19-information | https://news784.com |
| Suriname | https://covid-19.sr | https://www.diplomatie.gouv.fr | https://www.srherald.com |
| Trinidad and Tobago | http://www.news.gov.tt | https://tt.usembassy.gov | http://www.guardian.co.tt<br>https://trinidadexpress.com |
| Turks and Caicos Islands | https://www.gov.tc/moh/coronavirus/ | https://www.visittci.com/travel-info/turks-and-caicos-coronavirus-covid-19<br>https://www.gov.uk/foreign-travel-advice/turks-and-caicos-islands/staying-during-coronavirus | https://tcweeklynews.com |
| Germany | https://www.bundesregierung.de |  | https://www.thelocal.de |
| Iceland | https://www.government.is/government/covid-19 |  | https://en.wikipedia.org/wiki/COVID-19_pandemic_in_Iceland<br>https://www.landlaeknir.is<br>https://www.stjornarradid.is |
| Italy | http://www.governo.it<br>http://www.salute.gov.it<br>https://www.interno.gov.it | https://it.usembassy.gov/covid-19-information |  |
| Korea Republic of | http://www.mohw.go.kr | https://www.cdc.go.kr<br>https://kr.usembassy.gov/022420-covid-19-information |  |
| New Zealand | https://www.health.govt.nz<br>https://covid19.govt.nz<br>https://www.beehive.govt.nz |  | https://www.rnz.co.nz<br>https://www.nzherald.co.nz<br>https://www.newshub.co.nz |
| Singapore | https://www.moh.gov.sg<br>https://www.pmo.gov.sg<br>https://www.ecda.gov.sg<br>https://www.mom.gov.sg<br>https://www.moe.gov.sg |  | https://theaseanpost.com<br>https://en.wikipedia.org/wiki/COVID-19_pandemic_in_Singapore<br>https://www.straitstimes.com/singapore |
| Sweden | https://www.government.se<br>https://www.krisinformation.se |  |  |
| United Kingdom | www.legislation.gov.uk/ukxi/2020/327/pdfs/ukxi_20200327_en.pdf |  | https://www.bbc.com |
| Vietnam |  | https://vietnam.travel/things-to-do/information-travellers-novel-coronavirus-vietnam<br>https://vn.usembassy.gov/u-s-citizen-services/covid-19-information | http://news.chinhphu.vn<br>https://en.wikipedia.org/wiki/COVID-19_pandemic_in_Vietnam |

International media sources included:

https://www.bbc.com

https://www.bloomberg.com

https://apnews.com

https://www.theguardian.com

https://www.reuters.com

## Footnotes

1. Taxonomy of government measures adapted from Assessment Capacities Project (ACAPS) www.acaps.org/sites/acaps/files/resources/files/acaps_covid19_government_measures_dataset_0.xlsx)

1 ACAPS COVID-19 Government Measures Database. [Available from: https://www.acaps.org/covid19-government-measures-dataset.

